# Multidrug Antifungal Resistance and Clinical Outcomes in Fungal Keratitis: A Prospective Study in a South Indian Population

**DOI:** 10.64898/2026.02.02.26345336

**Authors:** Leonie Fingerhut, Ramakrishnan Vigneshwar, Florence Burté, Manojkumar Vimala Devi, Rathinavel Sethu Nagarajan, Rajarathinam Karpagam, Venkatesh Prajna, Bethany Mills, Prajna Lalitha

**Affiliations:** Institute for Regeneration and Repair, The University of Edinburgh, EH16 4UU, UK; Department of Cornea, Aravind Eye Care System, Madurai, 625 020, India; Department of Ocular Microbiology, Aravind Eye Care System, Madurai, 625 020, India

**Keywords:** Fungal keratitis, antifungal resistance, multidrug resistance, natamycin resistance, amphotericin B resistance, corneal infection

## Abstract

**Objectives:** Our primary objective was to identify the rate of anti-fungal resistance (including multidrug resistance) of fungal isolates cultured from fungal keratitis patients in South India. Our secondary objective was to identify associations between antifungal resistance and patient outcome.

**Subjects/Methods:** 153 patients attending the cornea clinic at Aravind Eye Hospital, Madurai (May – August 2025) with culture positive fungal keratitis were enrolled in the study. Cultured isolates were evaluated for natamycin, amphotericin B, voriconazole and econazole MIC (n=151-153). MIC data were analysed by Kruskal-Wallis test followed by Dunn’s multiple comparisons test. Patient disease outcome relative to isolate resistance was analysed by fisher’s exact test (n=91).

**Results:** There were high levels of resistance to all four antifungals, which were significantly elevated in *Fusarium* spp. and *Aspergillus* spp. isolates compared to other fungal genera (P<0.0001). Resistance of *Fusarium* spp. isolates (n= 81) to: natamycin: 38.3%; amphotericin B: 93.8%; voriconazole: 97.5% and econazole: 76.5%. Resistance of *Aspergillus spp.* isolates (n=33) to: natamycin: 66.7%; amphotericin B: 87.9%; voriconazole: 6.1%; and econazole: 0%. Overall, fungal resistance to natamycin significantly correlated with a poor clinical outcome, P=0.0034, relative risk 1.7 (95% CI: 1.2-2.6).

**Conclusions:** The majority of fungal isolates were resistant to multiple antifungals; none of the *Fusarium* isolates were susceptible to all four drugs, 15% were resistant to all of them. Resistance towards natamycin may worsen clinical outcome, particularly for infections caused by *Aspergillus*, even when susceptible to azoles. Our data supports the need for species specific MIC thresholds to have increased clinical utility.

## Introduction

Fungal keratitis remains a leading cause of visual impairment, blindness and eye-loss worldwide. It affects more than one million people annually, and corneal perforation is reported in up to 25% of cases (1, 2). While incidence is increasing across temperate climates, fungal keratitis disproportionately affects rural populations across the tropics, where the ophthalmic community has campaigned for neglected tropical disease status (3–7).

Fungal keratitis is challenging to treat, resulting in worse outcomes compared to bacterial keratitis (8, 9). Treatment relies on topical application of antifungal eye-drops, and options are limited, primarily to two drug classes: polyenes (e.g., natamycin, amphotericin B); and azoles (e.g., voriconazole, econazole) (10, 11). Natamycin prevails as the first-line treatment option, with inclusion of voriconazole or other drugs if there is no positive response to treatment (11, 12). Better outcomes with natamycin compared to voriconazole topical treatment have been demonstrated, particularly for treating *Fusarium* spp. infections (13).

This paucity of antifungal drugs to treat fungal keratitis may be compounded (now or in the future) by the increasing global incidence of antifungal resistance (14, 15). While a broad spectrum of fungi (393 species) have been cultured from fungal keratitis patients (16), 95% of cases are caused by either *Aspergillus spp.*, *Fusarium spp.*, or *Candida spp*.. Each of these genera have been categorised by the world health organisation (WHO) as critical and high priority pathogens (17). Antimicrobial-resistant *Aspergillus* spp. is further included in the 2019 U.S. Centers for Disease Control and Prevention watch list, and drug-resistant *Candida* species are listed as serious threats (18).

Antifungal susceptibility testing of fungi cultured from fungal keratitis patients is not routinely performed and/or reported, and therefore the scale of antifungal resistance is poorly understood. Moreover, how *in vitro* antifungal MIC values relate to fungal keratitis patient treatment outcomes is unknown. This hinders development of evidence-based policy and treatment guidelines.

Herein we report on the antifungal resistance profile of fungal isolates obtained from 153 fungal keratitis patients from South India between May – August 2025. The minimum inhibitory concentration (MIC) against each of the key clinically used antifungals (natamycin, amphotericin B, voriconazole, and econazole) for treatment of fungal keratitis were determined. Participant clinical symptoms were evaluated for up to one month to explore the relationship between antifungal resistance, clinical presentation and patient outcomes.

## Materials and methods

This prospective observational study was performed in the Cornea Clinic and Ocular Microbiology Laboratory of Aravind Eye Hospital, Madurai, India. The study was approved by the Institutional review board of Aravind Eye Hospital, Madurai (RET202500520).

The study included 153 consecutive culture-proven fungal keratitis cases from May to August 2025. Collection of patient clinical history, and standard clinical examination were performed. Visual acuity, location, size and features of the corneal ulcer were documented. These clinical details were captured at one-week and one-month follow-up appointments following the initial (baseline) visit, unless the patient did not attend the clinic and they were deemed “lost to follow-up”.

Corneal scraping specimens were collected at the first-appointment (at the time of study recruitment) under magnification by using a kimura spatula and evaluated with 10% potassium hydroxide wet mount and Gram stain. Subsequent scrapings were plated onto 5% sheep blood agar (incubated at 37°C) and potato dextrose agar (incubated at 25°C) and allowed to grow for at least seven days prior to testing. Any positive fungal isolate was identified to the genus or species level by Lactophenol Cotton Blue (LPCB) mount based on their spore morphology.

Antifungal susceptibility of filamentous fungi was determined using the broth microdilution method according to CLSI M38-A3 guidelines (19). The following antifungal agents were tested with the final concentration: Natamycin (0.06-32 µg/mL), amphotericin B (0.03-16 µg/mL), voriconazole (0.03-16 µg/mL), and econazole (0.03-16 µg/mL). The inoculum (conidial suspension) was prepared from 7-day-old cultures grown on potato dextrose agar and adjusted to a final inoculum of 0.4–5×10□ CFU mL^-1^ in sterile saline with 0.05% Tween 20. Natamycin, amphotericin B, voriconazole, and econazole were two-fold serially diluted in RPMI 1640 medium buffered with MOPS (pH 7.0). Each well of a 96-well microtiter plate was inoculated with 100 µL of fungal suspension and 100 µL of antifungal dilution, with growth and sterility controls included. The microtiter plates were incubated at 35°C for 48 to 72 hours. The minimum inhibitory concentration (MIC) was defined as the lowest drug concentration that resulted in complete (100%) inhibition of visible growth. Quality control was performed using reference strains.

MICs were interpreted according to CLSI breakpoints, epidemiological cut-off values where available. We used the following MIC breakpoints (20): Natamycin: ≥8.0□mg/L resistant; amphotericin B: ≥2.0□mg/L resistant; voriconazole: ≥2.0□mg/L resistant; and econazole: ≥8.0□mg/L resistant. Susceptible-increased exposure fungal isolates were treated as susceptible for further evaluation.

### Statistical analysis

Data were analysed using GraphPad Prism 10.4.2 (GraphPad Software, San Diego, CA, USA). For each antifungal tested, the mean antifungal MIC per fungi genera were compared using Kruskal-Wallis test followed by Dunn’s multiple comparisons test.

Spearman’s correlation was used to measure the strength and direction of association between clinical measure and log_2_ transformed MIC. Where applicable, participants who were lost to follow-up were excluded from analysis. Significance in correlation was considered when P<0.01. *R_s_*: ±0.00-0.19 = very weak correlation; ±0.2-0.39 = weak correlation; ±0.40-0.69 = moderate correlation; ±0.7-0.89 = strong correlation; and ±0.9-1.0 = very strong correlation. Significance for participant outcome against resistant/non-resistant fungal strain was determined by Fisher’s exact test, comparing healed vs not-healed outcome groups. *P ≤ 0.05; **P ≤ 0.01; ***P ≤ 0.001; and ****P ≤ 0.0001.

Upset plots were produced using the UpSetR package in R (version R 4.5.0) (21).

## Results

### Participant demographics and sample aetiology

A total of 153 fungal keratitis patients with culture-positive samples were included in this study. Participants’ ages ranged from seven to 88 years-old, with 59% identified as male (Supplementary Table S1). Half of the recruited patients had already received ocular medication when first presenting at the Aravind Eye Hospital, and had symptoms for a median time of five days. Samples from 81 patients grew *Fusarium* species (52.9%), from 33 patients *Aspergillus* species (21.6%), and 25.5% were other filamentous fungi. These included *Bipolaris spp.*, *Chaetomiaceae spp.*, *Colletotrichum spp.*, *Curvularia spp.*, *Exserohilum spp.*, *Lasiodiplodia spp.*, *Mucor spp.*, *Paecilomyces spp.*, *Papulospora equi*, *Pleiocarpon algeriense*, *Podospora spp.*, and *Scedosporium spp* (Supplementary Table S2).

### Antifungal minimum inhibitory concentrations (MICs) against cultured ocular clinical isolates

MICs were available for 151 fungal isolates against natamycin, amphotericin B, voriconazole and 150 against econazole. Across all isolates, the median MICs (interquartile range; IQR) were: 2.00 (1.00-8.00) µg/mL natamycin; 2.00 (0.50-8.00) µg/mL voriconazole; 2.00 (1.00-2.00) µg/mL amphotericin B; and 2.00 (0.50-8.00) µg/mL econazole. In total, 35.8% of the isolates were resistant to natamycin, 74.8% to amphotericin B, 58.3% to voriconazole and 41.7% to econazole.

Comparison of antifungal MICs for *Fusarium* and *Aspergillus* genera identified a different resistance pattern between them and the combined other genera (Figure 1, Supplementary Figure S1).

**Figure 1.**
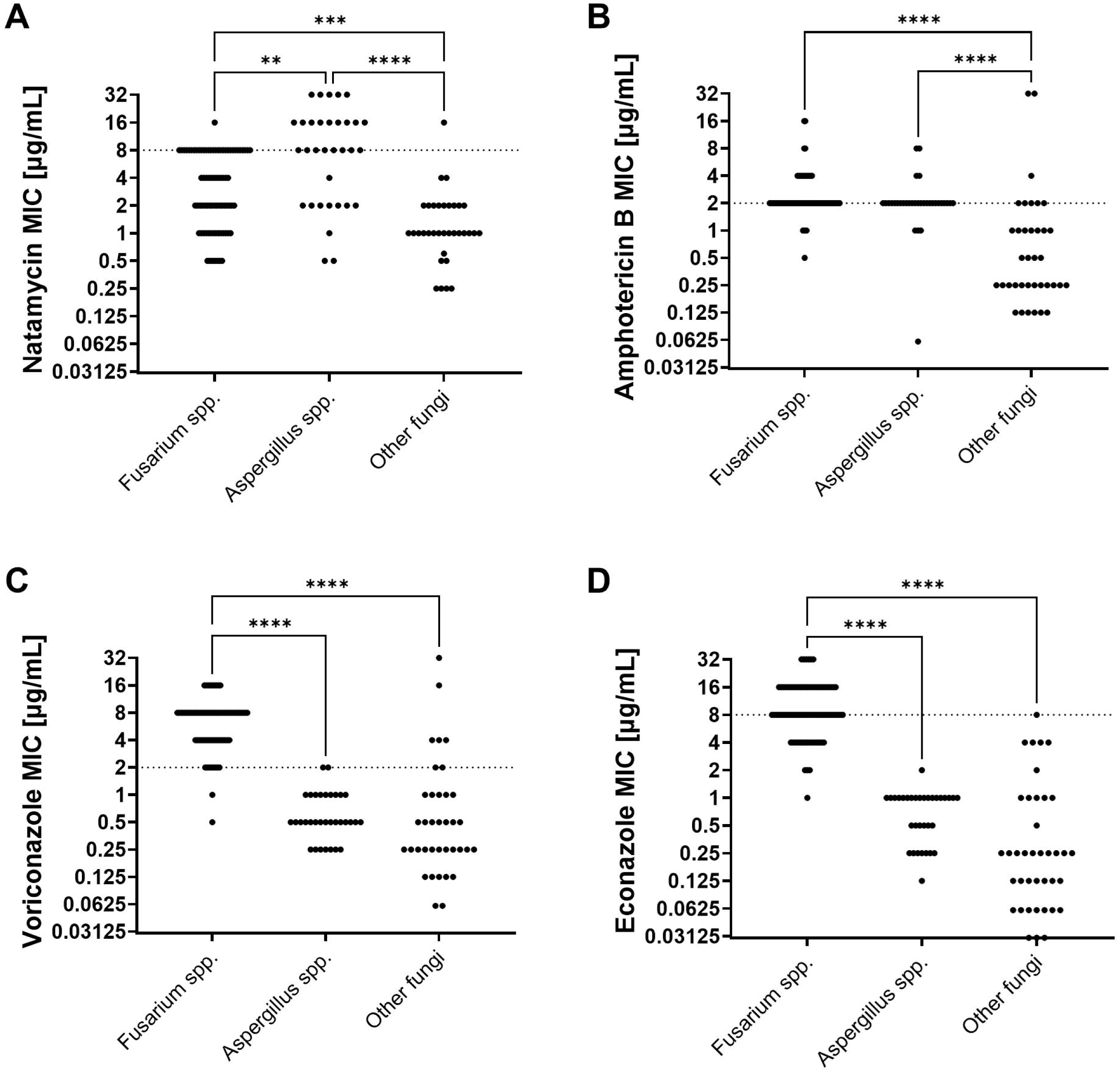
Minimum inhibitory concentrations (MICs) of antifungals against fungal keratitis isolates. MIC [µg/mL] of the antifungals a) natamycin, b) amphotericin B, c) voriconazole and d) econazole against fungal isolates cultured from fungal keratitis patients. Cut-off values for resistance are 8 µg/mL for natamycin and econazole, and 2 µg/mL for voriconazole and amphotericin B (depicted by horizontal line). Statistical analysis was performed by Kruskal-Wallis test followed by Dunn’s multiple comparisons test (**p<0.01, ***p<0.001, ****p<0.0001). Each data point represents one isolate.

The isolates of *Fusarium* spp. showed resistance against natamycin in 38.9% of cases, against amphotericin B in 93.5% of cases, against voriconazole in 97.5% of cases, and 76.5% of the isolates were resistant to econazole.

While the *Aspergillus spp.* isolates demonstrated high levels of resistance towards natamycin (66.7% of isolates) and amphotericin B (87.9% of isolates), only 6.1% of isolates were resistant to voriconazole, and none were resistant to econazole.

### Fungal keratitis ocular isolate resistance to multiple antifungal drugs

Just 27 isolates were susceptible to all four of the antifungal drugs tested, none of those isolates being *Fusarium* spp., and only one *Aspergillus flavus* isolate. The majority (66.9%) of the ocular isolates were resistant to more than one of the antifungals. Furthermore, 23 (15.2%) *Fusarium* spp. isolates were resistant to all four antifungals (Figure 2).

**Figure 2.**
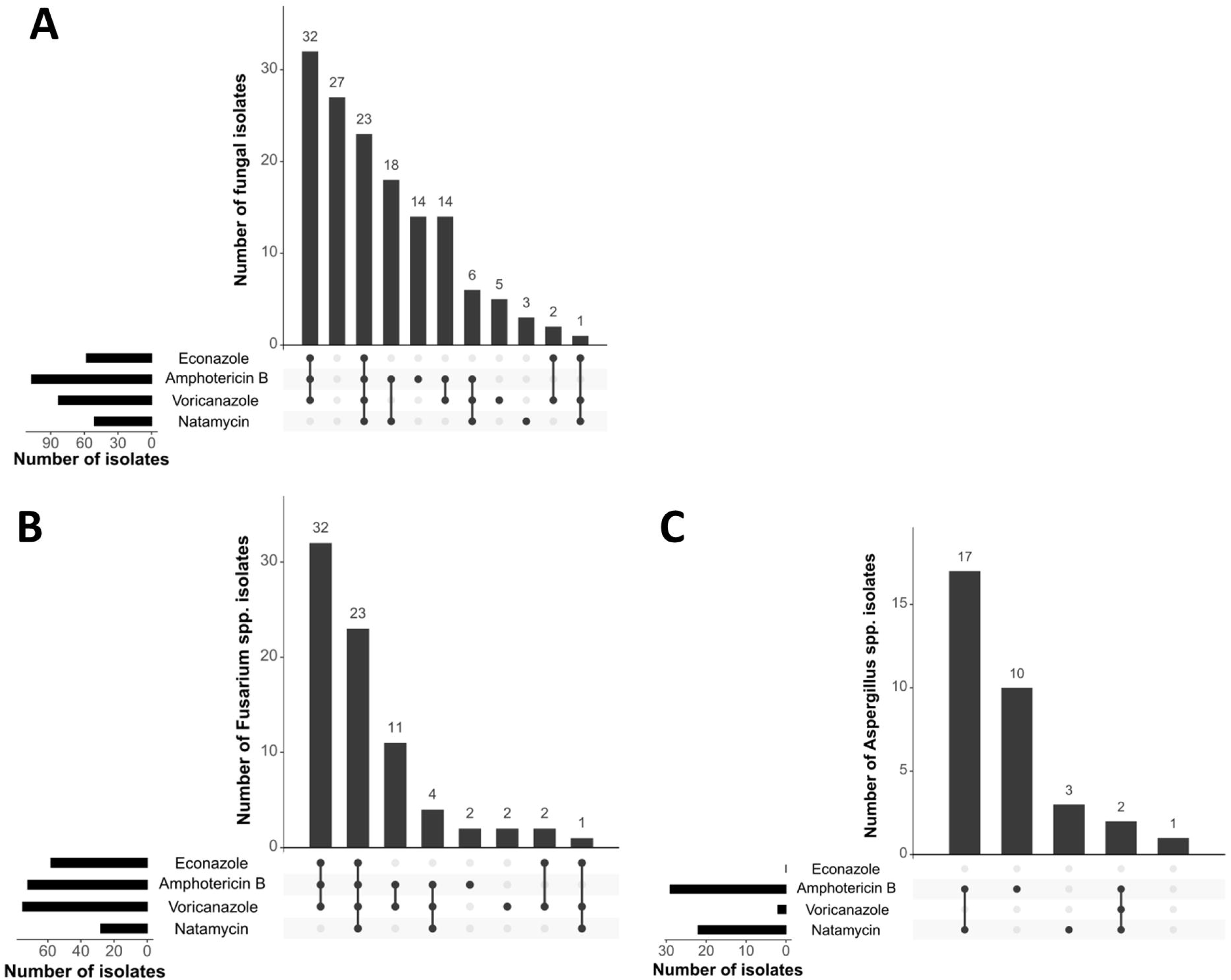
Concurrent resistance of fungal keratitis isolates to antifungal drugs. Upset plots depicting the number of a) all fungal isolates; b) Fusarium spp.; and c) Aspergillus spp. isolates with resistance to common antifungal drugs. MIC cut-off values for resistance are 8 µg/mL for natamycin and econazole, and 2 µg/mL for voriconazole and amphotericin B. Black dots indicate resistance.

Spearman’s rank correlation analysis identified a very strong positive correlation between voriconazole and econazole MIC, and moderate positive correlation between amphotericin B MIC and each of the azoles (P<0.0001) (Table 1).

**Table 1:** Correlation between fungal isolate MICs.

| **Table 1: Correlation between fungal isolate MICs** |  |  |  |  |  |
| --- | --- | --- | --- | --- | --- |
| **MIC** | **r<sub>s</sub>** | **95% CI** | **Strength of correlation** | **P** | **(n)** |
| Natamycin vs Amphotericin B | 0.30 | 0.14 to 0.44 | weak | 0.0002 | 151 |
| Natamycin vs Voriconazole | 0.21 | 0.052 to 0.37 | weak | 0.0082 | 151 |
| Natamycin vs Econazole | 0.17 | 0.002 to 0.32 | very weak | 0.0413 | 150 |
| Amphotericin B vs Voriconazole | 0.49 | 0.35 to 0.60 | moderate | <0.0001 | 151 |
| Amphotericin B vs Econazole | 0.53 | 0.39 to 0.64 | moderate | <0.0001 | 150 |
| Voriconazole vs Econazole | 0.87 | 0.82 to 0.90 | very strong | <0.0001 | 150 |
| Trends determined by Spearman's rank correlation coefficient. Positive r<sub>s</sub> indicates positive correlation. Significance considered when P<0.01 |  |  |  |  |  |

### Prior use of antifungals and fungal isolate sensitivity

Many of the patients (n=74, 48%) had taken some form of topical treatment for their corneal ulcer prior to enrolment into the study. 62 (84%) of these participants had dosed at least one antibiotic, 28 (32%) had dosed at least one azole and 39 (53%) had dosed one polyene (38/39 = natamycin). 23 (31%) had previously been treated with a combination of one or more azoles and a polyene. The majority of patients with an antifungal resistant infection had no history of taking the corresponding anti-fungal: 64.8% of patients with natamycin resistant fungi had no history of natamycin use; 99.1% of patients with amphotericin B resistance had no history of using it, similarly, 89.5% and 93.2% of patients with fungal isolates resistant to voriconazole and econazole had no history of using these antifungal drugs, respectively. However, prior use of natamycin or econazole was found to increase the relative risk of resistance by 1.7-fold (95% CI: 1.1-2.5) and 2.6-fold respectively (95% CI:1.3-6.4) (Supplementary Table S3).

### Antifungal MIC: Clinical presentation and progression

The study participants underwent clinical assessment at the time of enrolment (baseline), at 1 week and at 1 month. In total, 91 (59.5%) patients had a known outcome: 59 patients healed and 32 had poor outcomes (i.e. required therapeutic penetrating keratoplasty, TPK). In total, 62 (40.5%) patients were lost to follow-up and their outcome is unknown (Supplementary Figure S2).

Key clinical measures (visual acuity, epithelial defect area, stromal infiltrate depth, and ulcer severity score) are reported in Supplementary Table S4 and Supplementary Figure S3. Epithelial defects significantly improved across the cohort within one week (P<0.0001). Ulcer severity did not improve between baseline and week one, however this was significantly improved for patients remaining in the study at one month (P<0.0001).

There was no association between patient sex or admission route (free vs paying) and MIC, however there were weak, but significant trends between patient age and voriconazole (P=0.0003) and econazole MICs (P=0.0002), with higher MICs being associated with younger patient ages. At presentation (baseline), higher natamycin MIC very weakly correlated with a worse ulcer severity score (p=0.0016) (Supplementary Table S5).

### Antifungal resistance: Clinical outcome

Once entered into the study, patient treatment plans were similar across the study cohort (Supplementary Figure S4). 80.4% of patients were prescribed natamycin plus one or more azole (healed: 81.4%; poor-outcome: 87.5%; and lost to follow-up: 77.4%), with econazole the most frequently prescribed azole. Lost to follow-up patients were excluded from study outcome analysis.

Natamycin resistance (MIC≥ 8 µg/mL) significantly affected the patient outcome (P=0.0034, Table 2), with attributable risk of a poor outcome increased by 31% (95% CI: 9% to 50%). Species sub-analysis determined that this relationship was genera dependent, and was significant for the 33 cases caused by *Aspergillus* spp. (P=0.0251, Table 2), but not for keratitis caused by *Fusarium* spp..

**Table 2.**
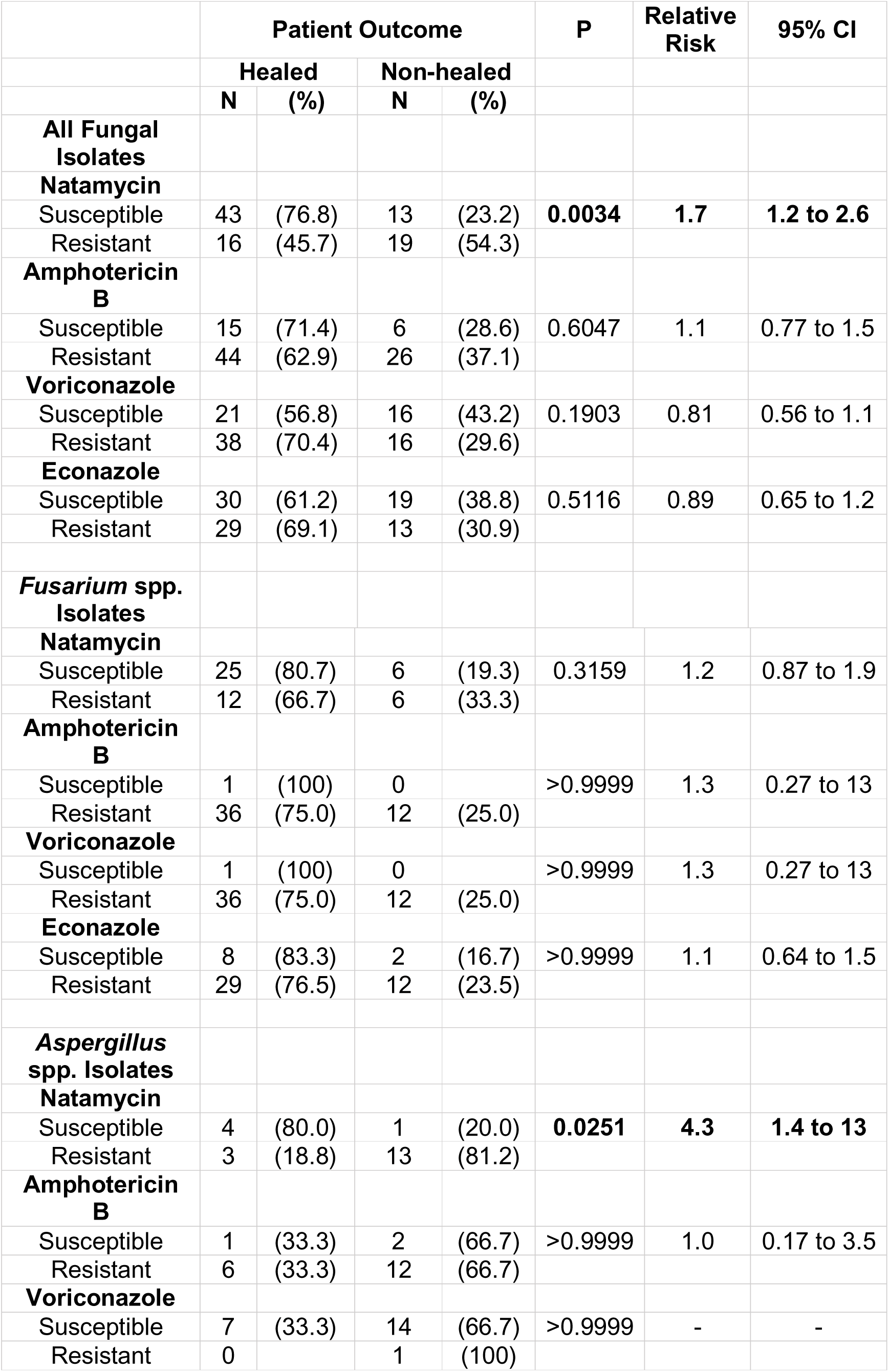
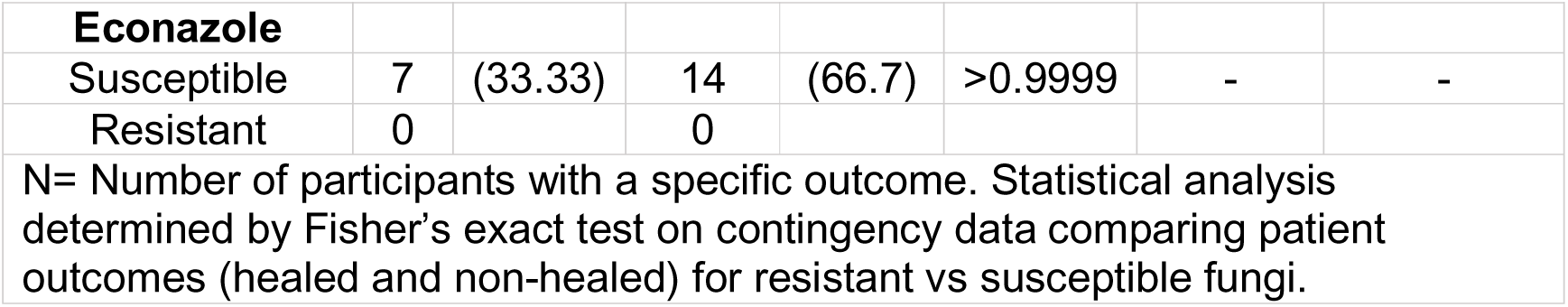
Patient outcome in relation to anti-fungal resistance.

## Discussion

Antifungal resistance is of increasing concern due to the paucity of antifungal treatments available. Increasing trends have been reported worldwide, often attributed to increased use of fungicides (azoles) in agriculture and industry, and improper medical application (22). The majority of fungi in our study were resistant to two or more antifungals, including those from both polyene and azole classes. While prior use of antibiotics has been shown to increase the MIC of ocular bacterial isolates(23) the majority of patients with resistant strains in this study had no history of taking the corresponding antifungal. This perhaps indicates that the selection pressure driving increased MICs for these isolates is occurring within the environment. Azoles are frequently used in agriculture, and concerns about the contribution of these practices to driving azole resistance are well documented (22, 24). The majority of patients attending the cornea clinic in this study are agriculture workers, and/or suffered cornea injury via vegetative matter, providing a direct route of transmission for these opportunistic pathogens from environments under high selection pressure.

Notably, while azole resistant *Aspergillus* spp. is most commonly reported in the literature as a consequence of their use in agriculture, only two (6%) of the Aspergillus isolates in this study were resistant to voriconazole, and none were resistant to econazole. This is in contrast to *Fusarium* spp, where 97% of isolates were resistant to voriconazole, and 75% were resistant to econazole. It is less clear what is driving selection pressure of fungal resistance towards amphotericin B (94% of Fusarium isolates and 88% of Aspergillus isolates) and natamycin (39% Fusarium isolates and 67% Aspergillus isolates) and warrants further investigation through a One Health approach to examine the environment-human transmission route(24, 25).

Surveillance of present and emerging antimicrobial resistance is imperative to this agenda, and for devising local, evidence-based policies around stewardship and clinical prescribing. Despite its importance, few studies have reported the antifungal resistance profile of isolates cultured from fungal keratitis patients. Where resistance is reported, differences in resistance breakpoints among studies make comparisons challenging (12, 26–28). However, a recent 585 patient study from the UK (using the same resistance break-points as used within our study) determined antifungal MICs of corneal and contact lens isolates(20). While our study found a similar proportion of fungal isolates resistant to econazole, a substantially higher proportion of our south Indian isolates showed resistance to natamycin (36% vs 8.2%), amphotericin B (74.8% vs 27.2%) and voriconazole (58.3% vs 27%). This was more pronounced when comparing data from *Fusarium* spp., and *Aspergillus* spp. isolates across each of the antimicrobials. For example, none of the UK *Fusarium* spp. isolates showed resistance to natamycin, compared to almost 40% of our isolates. Our study identified that 94% of *Fusarium* spp. isolates were resistant to amphotericin B, compared to just one-third in the UK study, and 88% of the *Aspergillus* spp. isolates in our study were resistant to amphotericin B compared to just 3% in the UK study. These differences echo geographical differences also observed for antimicrobial resistance against bacterial isolates and highlight the need for region-specific antimicrobial resistance surveillance.

A secondary objective of our study was to examine antifungal resistance/susceptibility in relation to clinical outcomes. While our sample sizes are relatively small due to patient loss to follow-up, for those remaining in the study to a final outcome (n=91), natamycin resistance significantly increased the relative risk of a poor outcome (i.e. TPK), compared to those which were susceptible, and this was most pronounced for cases caused by *Aspergillus* spp.. However, our analysis also determined that higher natamycin MIC correlated with increased time to seek healthcare and a worse ulcer severity score at the time of presentation – both known risk factors for poor outcomes (2). Whether natamycin resistance is truly causative of worse clinical outcomes is unclear (29, 30), and warrants further investigation in adequately powered studies with standardised methodologies.

As already discussed, few fungal keratitis antifungal resistance studies have been reported, and even fewer have reported concurrent multidrug resistance profiles and/or how resistance may affect patient outcomes. This is a particular strength of our study. A limitation of ours and other studies is the lack of standardised, clinically appropriate MIC breakpoints for fungal keratitis. MICs are typically based on systemic drug concentrations, rather than topical application, where the local concentration of the drug may be higher. Furthermore, they do not consider the increased MIC required to treat fungal biofilms, which can be prevalent in fungal keratitis cases (4, 31), nor species-specific differences in breakpoints. For example, based on our study breakpoints, 13 out of 14 “poor outcome” *Aspergillus* spp. patients were treated with an “appropriate” azole based on the susceptibility profile, whereas 25% of patients with *Fusarium* infections successfully healed, despite having isolates resistant to all of their prescribed drugs, based on *in vitro* MIC values.

A systematic review on the topic is warranted to consolidate the literature and identify key areas of uncertainty and contribute to standardised reporting methodology, and disease and species-specific MIC breakpoints. Interpreting antifungal MIC and clinical consequences of resistance are imperative to the stewardship of existing antimicrobials and the development of new therapeutic strategies.

## Data Availability

All data produced in the present study are available upon reasonable request to the authors

## Acknowledgements

BM, LF and FB were supported by a UKRI Future Leaders Fellowship (United Kingdom, No. MR/V026097/1). There are no further financial disclosures or conflicts of interest.

## Author Contributions (CRediT)

LF: Formal analysis, Data curation, Writing – original draft; RV: Data curation, Investigation, Writing – review and editing; FB: Formal analysis, Writing – original draft; MVD: Data curation, Investigation; RSN: Data curation, Investigation; RK: Data curation, Investigation; VP: Conceptualization, Data curation, Methodology, Supervision, Writing – review and editing; BM: Conceptualization; Funding acquisition, Data curation, Methodology, Supervision, Writing – original draft & PL: Conceptualization, Data curation, Methodology, Supervision, Writing – review and editing.

## Data availability statement

Data are available on reasonable request. Supplementary information is available at Eye’s website.

For the purpose of open access, the author has applied a Creative Commons Attribution (CC BY) licence to any Author Accepted Manuscript version arising from this submission.

## Ethics Statement

The study was approved by the Institutional Review Board of Aravind Eye Hospital, Madurai (RET202500520).

