## Supplemental Figures and Tables for "Multidrug Antifungal Resistance and Clinical Outcomes in Fungal Keratitis: A Prospective Study in a South Indian Population"

| Supplementary Table 1. Baseline patient characteristics and final patient outcome | | | | | | |
|---|---|---|---|---|---|---|
| | N[a] | | N[b] | (%) | Median | (IQR) |
| **Sex** | 153 | Male | 90 | (58.8) | | |
| | | Female | 63 | (41.2) | | |
| **Age** | 153 | Years | | | 51 | (37-63) |
| **Admission route** | 153 | Free | 89 | (58.2) | | |
| | | Paying | 64 | (41.8) | | |
| **Durations of symptoms** | 149 | Days | | | 5 | (3-7) |
| **Prior ocular medication** | 153 | Yes | 77 | (50.3) | | |
| | | No | 76 | (49.7) | | |
| **Visual acuity** | 153 | LogMAR | | | 1 | (0.32-2.00) |
| **Epithelial defect** | 149 | $mm^2$ | | | 7.84 | (1.5-16.0) |
| **Stromal infiltrate depth** | 132 | % | | | 40 | (30-70) |
| **Ulcer severity score*** | 151 | 0-3 | | | 2 | (2-2) |
| **Fungal species cultured** | 153 | *Fusarium* spp. | 81 | (52.9) | | |
| | | *Aspergillus* spp. | 33 | (21.6) | | |
| | | Other | 39 | (25.5) | | |
| **Final Outcome** | 153 | Healed | 59 | (38.6) | | |
| | | Non-healed | 32 | (20.9) | | |
| | | Lost to follow-up | 62 | (40.5) | | |

**N[a] = number of participants with data available. N[b] = number or participants per category. IQR = Interquartile range. *Severity score was recorded as: 0 = healed, 1 = mild, 2 = moderate, 3 = severe.**

| **Supplementary Table 2. Fungal species cultured from corneal scrapes from fungal keratitis patients.** | | |
|---|---|---|
| **Fungal Species** | **Number of isolates** | **(%)** |
| *Fusarium spp.* | 81 | (52.94) |
| *Aspergillus flavus* | 25 | (16.34) |
| *Aspergillus fumigatus* | 5 | (3.27) |
| *Aspergillus terreus* | 3 | (1.96) |
| *Curvularia spp.* | 11 | (7.19) |
| *Lasiodiplodia spp.* | 8 | (5.23) |
| *Bipolaris spp.* | 5 | (3.27) |
| *Exserohilum spp.* | 4 | (2.61) |
| *Scedosporium spp.* | 3 | (1.96) |
| *Papulospora equi* | 2 | (1.31) |
| *Chaetomiaceae sp.* | 1 | (0.65) |
| *Colletotrichum gloesporioides* | 1 | (0.65) |
| *Mucor sp.* | 1 | (0.65) |
| *Paecilomyces sp.* | 1 | (0.65) |
| *Pleiocarpon algeriense* | 1 | (0.65) |
| *Podospora sp.* | 1 | (0.65) |

**Supplementary Table 3. Antifungal resistance of clinical isolate in relation to the same drug taken prior to study enrolment**

| **Antifungal evaluated[#]** | | **N[a]** | **(%)** | **N[b]** | **(%)** | **P** | **Relative Risk** | **95% CI** |
|---|---|---|---|---|---|---|---|---|
| **Natamycin** (n=151) | Susceptible | 18 | (18.6) | 79 | (81.4) | **0.0299** | **1.7** | 1.1 to 2.5 |
| | Resistant | 19 | (35.2) | 35 | (64.8) | | | |
| **Amphotericin B** (n=149) | Susceptible | 0 | | 37 | (100) | >0.9999 | 1.3 | 0.27 to 2.9 |
| | Resistant | 1 | (0.9) | 111 | (99.1) | | | |
| **Voriconazole** (n=148) | Susceptible | 10 | (16.1) | 52 | (83.9) | 0.3295 | 0.79 | 0.45 to 1.2 |
| | Resistant | 9 | (10.5) | 77 | (89.5) | | | |
| **Econazole** (n=144) | Susceptible | 0 | | 86 | (100) | **0.0247** | **2.6** | 1.3 to 6.4 |
| | Resistant | 4 | (6.8) | 54 | (93.2) | | | |

N[a] = number of participants who had taken the drug indicated in column # prior to diagnostic scraping & specimen identification. N[b] = number of participants who had not taken the drug indicated in column # prior to diagnostic scraping & specimen identification - determined from patient self-reported history obtained at baseline. Statistical analysis determined by Fisher’s exact test on contingency data comparing isolate susceptibility/resistance to antifungal treatment prior to study enrolment.

| Supplementary Table 4. Baseline and follow-up visit patient data. | | | | | | | | | |
|---|---|---|---|---|---|---|---|---|---|
| | Baseline | | | 1 week | | | 1 month | | |
| | N | Median | IQR | N | Median | IQR | N | Median | IQR |
| **Visual acuity (LogMAR)** | 153 | 1 | 0.32-2.0 | 120 | 1 | 0.48-1.96 | 83 | 1.1 | 0.48-2.3 |
| **Epithelial defect area ($mm^2$)** | 149 | 7.84 | 1.5-16.0 | 108 | 1 | 0-8.71 | 77 | 0 | 0-0 |
| **Stromal infiltrate depth (%)** | 132 | 40 | 30-70 | 87 | 40 | 30-70 | 49 | 35 | 0-80 |
| **Ulcer severity score*** | 151 | 2 | 2-2 | 114 | 2 | 2-2 | 81 | 1 | 0-2 |

N= number of participants with data. IQR = interquartile range. *Severity score was recorded as: 0 = healed, 1 = mild, 2 = moderate, 3 = severe.

| **Supplementary Table 5. Correlation clinical feature vs fungal isolate MIC at study baseline** | | | | | |
|---|---|---|---|---|---|
| | **$r_s$** | **95% CI** | **Strength of correlation** | **P** | **(n)** |
| **Age** | | | | | |
| Natamycin MIC | -0.07 | -0.23 to 0.095 | very weak | 0.3882 | 151 |
| Amphotericin B MIC | -0.20 | -0.36 to -0.040 | weak | 0.0126 | 151 |
| Voriconazole MIC | **-0.29** | **-0.43 to -0.13** | **weak** | **0.0003** | 151 |
| Econazole MIC | **-0.30** | **-0.44 to -0.14** | **weak** | **0.0002** | 150 |
| **Duration of symptoms (days) to first seek help** | | | | | |
| Natamycin MIC | **0.26** | **0.10 to 0.41** | **weak** | **0.0012** | 149 |
| Amphotericin B MIC | 0.01 | -0.16 to 0.17 | very weak | 0.9382 | 149 |
| Voriconazole MIC | -0.05 | -0.21 to 0.12 | very weak | 0.5510 | 149 |
| Econazole MIC | -0.10 | -0.26 to 0.066 | very weak | 0.2205 | 148 |
| **Duration of symptoms (days) to AEH** | | | | | |
| Natamycin MIC | 0.21 | 0.043 to 0.36 | weak | 0.0114 | 147 |
| Amphotericin B MIC | -0.01 | -0.18 to 0.16 | very weak | 0.9088 | 147 |
| Voriconazole MIC | -0.10 | -0.26 to 0.066 | very weak | 0.2209 | 147 |
| Econazole MIC | -0.18 | -0.34 to -0.016 | very weak | 0.0270 | 146 |
| **LogMAR** | | | | | |
| Natamycin MIC | 0.18 | 0.015 to 0.33 | very weak | 0.0282 | 151 |
| Amphotericin B MIC | 0.03 | -0.13 to 0.20 | very weak | 0.6758 | 151 |
| Voriconazole MIC | -0.10 | -0.26 to 0.064 | very weak | 0.2165 | 151 |
| Econazole MIC | -0.15 | -0.30 to 0.020 | very weak | 0.0764 | 150 |
| **Epithelial defect area ($mm^2$)** | | | | | |
| Natamycin MIC | 0.13 | -0.041 to 0.29 | very weak | 0.1262 | 147 |
| Amphotericin B MIC | 0.12 | -0.046 to 0.28 | very weak | 0.1424 | 147 |
| Voriconazole MIC | 0.06 | -0.11 to 0.22 | very weak | 0.4956 | 147 |
| Econazole MIC | 0.01 | -0.16 to 0.17 | very weak | 0.9294 | 146 |
| **Stromal infiltrate depth (%)** | | | | | |
| Natamycin MIC | -0.03 | -0.20 to 0.15 | very weak | 0.7627 | 130 |
| Amphotericin B MIC | 0.01 | -0.17 to 0.19 | very weak | 0.8895 | 130 |
| Voriconazole MIC | -0.09 | -0.26 to 0.092 | very weak | 0.3257 | 130 |
| Econazole MIC | -0.11 | -0.29 to 0.066 | very weak | 0.2024 | 129 |

| **Supplementary Table 5. Correlation clinical feature vs fungal isolate MIC at study baseline (cont.)** | | | | | |
|---|---|---|---|---|---|
| | **$r_s$** | **95% CI** | **Strength of correlation** | **P** | (n) |
| **Severity score** | | | | | |
| Natamycin MIC | **0.26** | **0.094 to 0.40** | **weak** | **0.0016** | 149 |
| Amphotericin B MIC | 0.15 | -0.019 to 0.30 | very weak | 0.0743 | 149 |
| Voriconazole MIC | 0.00 | -0.17 to 0.16 | very weak | 0.9923 | 149 |
| Econazole MIC | -0.06 | -0.22 to 0.11 | very weak | 0.4764 | 148 |
| Trends determined by Spearman's rank correlation coefficient. Negative $r_s$ indicates negative corelation, positive $r_s$ indicates positive correlation. Significance considered when P<0.01 | | | | | |

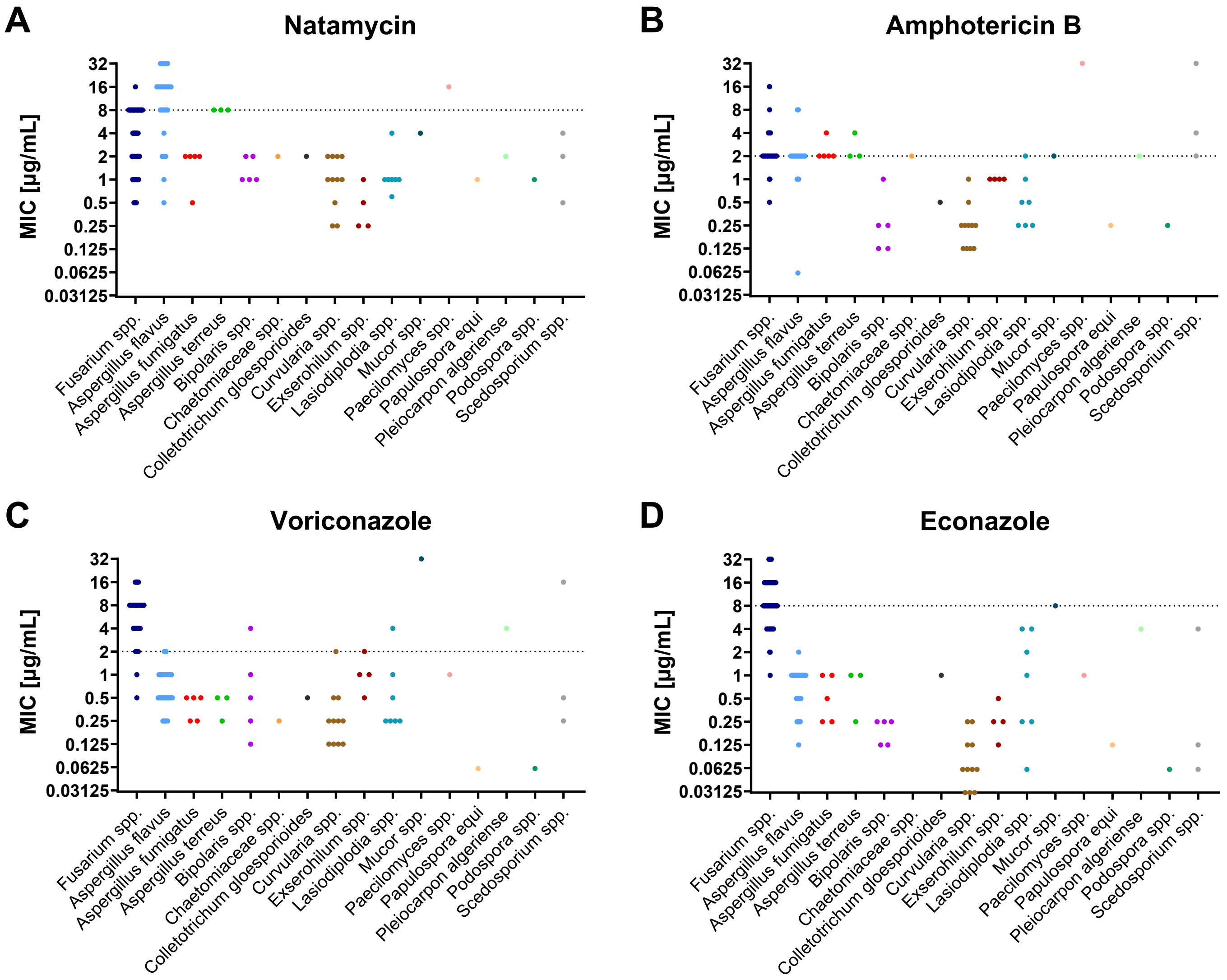

**Supplementary Figure 1. Minimum inhibitory concentrations (MICs) of antifungals against fungal keratitis isolates.**

MIC [µg/mL] of the antifungals **a)** natamycin, **b)** amphotericin B, **c)** voriconazole and **d)** econazole against fungal isolates cultured from fungal keratitis patients. Cut-off values for resistance are 8 µg/mL for natamycin and econazole, and 2 µg/mL for voriconazole and amphotericin B (depicted by horizontal line). Each data point represents one isolate.

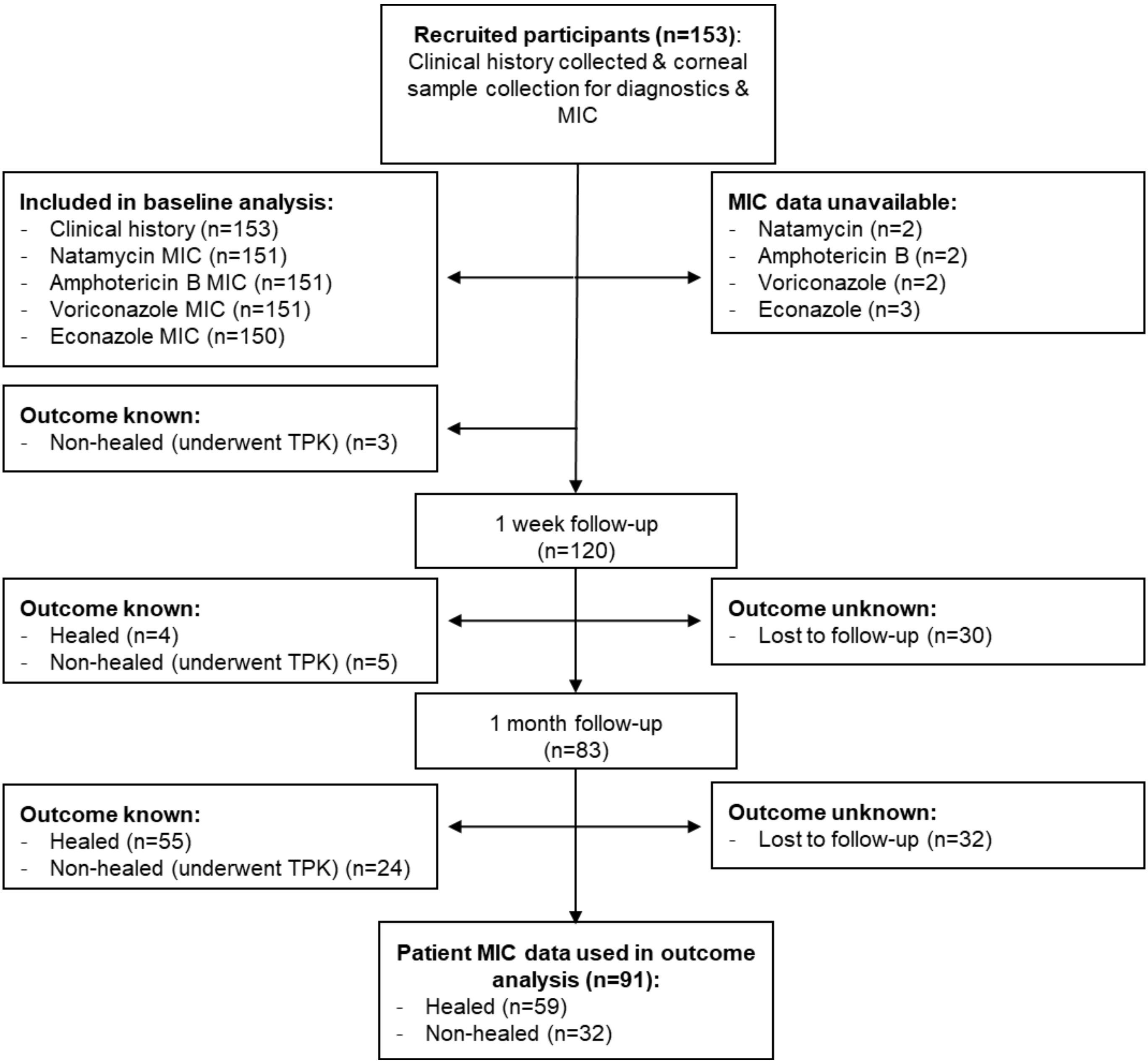

**Supplementary Figure 2. Schematic of study participant flow over the study duration.**

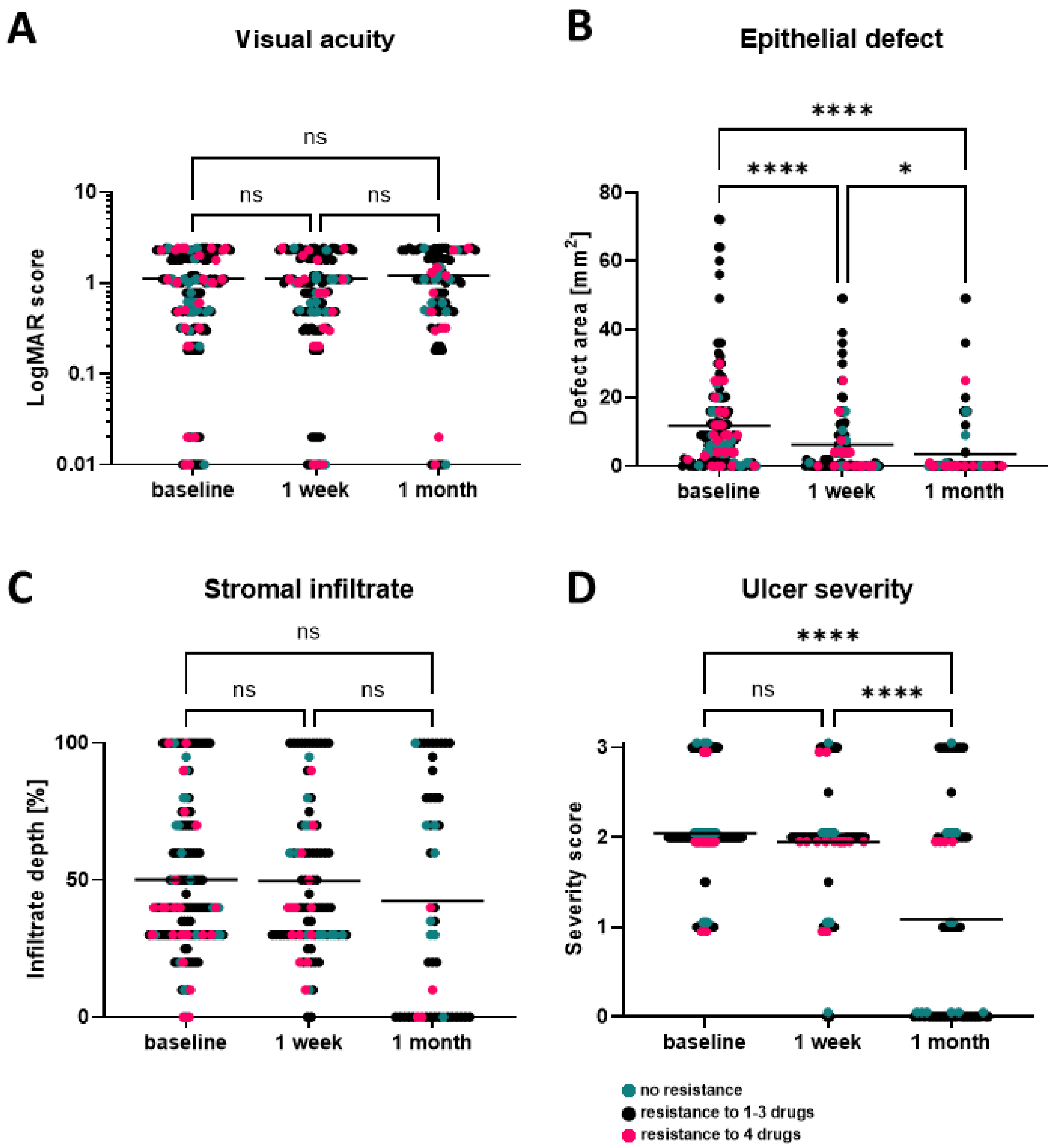

**Supplementary Figure 3. Clinical symptoms of fungal keratitis patients over time.**

Clinical measures of fungal keratitis patients were recorded upon enrolment in the study (baseline), at 1-week and 1-month follow-up examinations. Of these, **a)** visual acuity, **b)** epithelial defect size, **c)** stromal infiltrate depth and **d)** ulcer severity score (0 = healed, 1 = mild, 2 = moderate, 3 = severe) are displayed, with color-coded resistance level. Green values are from patients with susceptibility to all four tested antifungals; isolates from patients in black had one to three resistances. Red displays patient data with isolates resistant against all four antifungals. Data points of ulcer severity score were slightly shifted for display purposes. Statistical analysis was performed with mixed-effect analysis, followed by Tukey's multiple comparisons test (*p<0.05, ****p<0.0001). Each data point represents one patient.

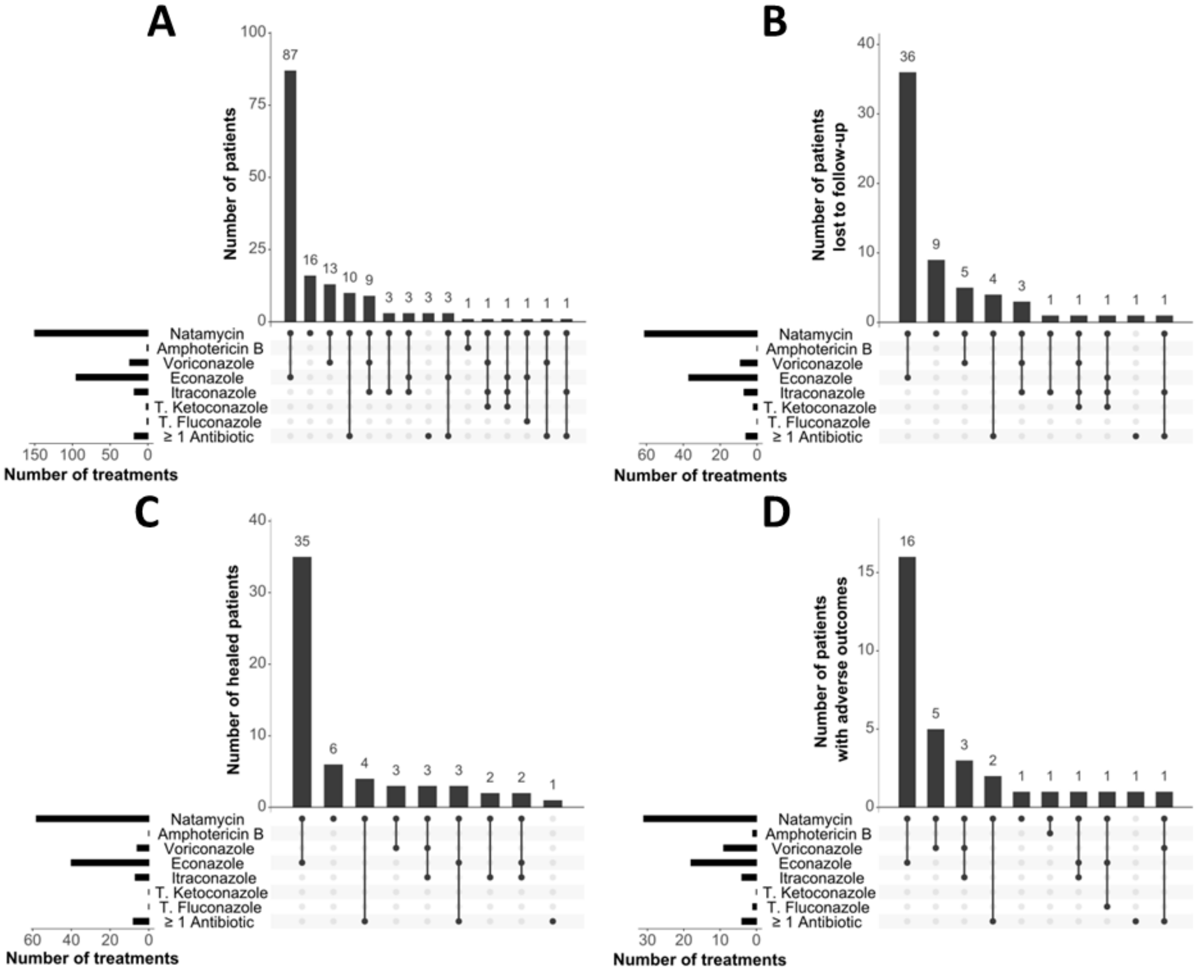

**Supplementary Figure 4. Prescribed treatments for fungal keratitis patients enrolled in the study.** Upset plots depicting the number of a) all patients; b) patients lost to follow-up; c) patients who healed; d) patients who had an adverse outcome. Black dots indicate prescribed drug. Natamycin, amphotericin B, voriconazole, econazole, itraconazole and antibiotics prescribed as topical eye-drops. Ketoconazole and fluconazole prescribed as oral tablets.

.
